# Differential Outcomes with Empagliflozin and Dapagliflozin in Heart Failure with Mildly Reduced Ejection Fraction

**DOI:** 10.64898/2026.01.09.26343819

**Authors:** Ibrahim Mortada, Aaron Lee, Ayushi Sahu, Erin Vidal, Krishna Paul, Yazeed Alhanbali, Shareef Mansour, Khaled Chatila, Dietrich Jehle, Thomas Blackwell, Hani Jneid

## Abstract

**Background:** Sodium–glucose cotransporter-2 (SGLT2) inhibitors improve outcomes in heart failure (HF), yet comparative effectiveness between individual agents in heart failure with mildly reduced ejection fraction (HFmrEF) remains limited.

**Methods:** We conducted a retrospective cohort study using the TriNetX Global Collaborative Network electronic health record database. Adults (≥18 years) with HF and left ventricular ejection fraction (LVEF) 41–49% initiating empagliflozin or dapagliflozin between September 2021 and December 2025 were included. Propensity score matching (1:1) balanced demographics, comorbidities, medications, and laboratory values, yielding 1,466 patients per group. Outcomes were assessed over 1 year after treatment initiation and included all-cause mortality, hospitalization, HF exacerbation, and urinary tract infection (UTI). Risk analyses and Kaplan–Meier survival analyses with hazard ratios (HRs) were performed.

**Results:** In the matched cohort, empagliflozin was associated with lower all-cause mortality (HR 0.75, 95% CI 0.59–0.96), substantially fewer hospitalizations (HR 0.61, 95% CI 0.55–0.67), and HF exacerbations (HR 0.64, 95% CI 0.57–0.73) compared to dapagliflozin. Rates of UTI were similar between groups (HR 0.81, 95% CI 0.65–1.01).

**Conclusions:** In this large real-world HFmrEF population, empagliflozin was associated with lower mortality, hospitalization, and HF exacerbation compared with dapagliflozin, with no significant difference in UTI risk. These findings suggest potential heterogeneity in clinical effectiveness among SGLT2 inhibitors in HFmrEF and warrant confirmation in prospective comparative studies.

**Graphical Abstract:** 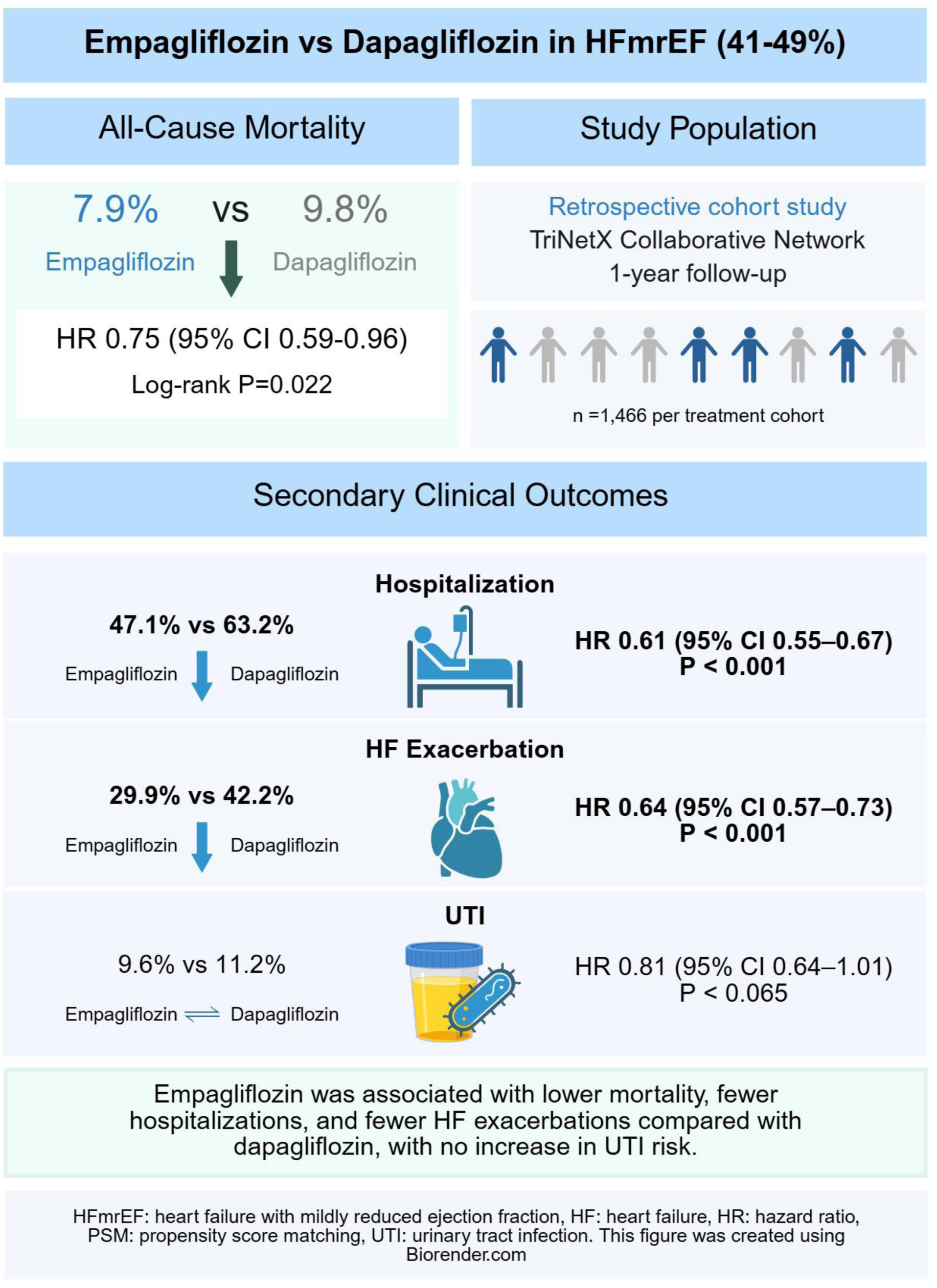

## Introduction

Sodium-glucose cotransporter-2 (SGLT2) inhibitors, including empagliflozin and dapagliflozin, have been shown to reduce cardiovascular mortality and heart failure (HF) related hospitalizations in patients with HF ^1–3^. However, therapeutic effects are not necessarily uniform across medications within the same pharmacologic class. Emerging evidence suggests empagliflozin may confer stronger benefits in heart failure with reduced ejection fraction (HFrEF)^4^, while dapagliflozin could offer enhanced efficacy in heart failure with preserved ejection fraction (HFpEF)^5,6^. Despite these observations, data directly comparing SGLT2 inhibitors across the spectrum of HF phenotypes remain limited. In particular, heart failure with mildly reduced ejection fraction (HFmrEF) represents an understudied and therapeutically heterogeneous population, for whom evidence guiding agent-specific SGLT2 inhibitor selection is sparse. A 2024 real-world analysis compared empagliflozin and dapagliflozin across broad EF categories but dichotomized patients into preserved and reduced EF groups, without specifically isolating the midrange phenotype^3^. Similarly, a multicenter cohort study reported comparable outcomes between empagliflozin and dapagliflozin across EF strata, including midrange EF, within a different healthcare setting ^7^. However, HFmrEF represents a clinically distinct and heterogeneous population that remains underrepresented in comparative analyses. Accordingly, the present study was designed to evaluate the comparative effectiveness of empagliflozin versus dapagliflozin on clinical outcomes, specifically among patients with HFmrEF.

## Methods

TriNetX is a global federated health research network that provides de-identified electronic medical records (EMRs) across participating healthcare organizations (HCOs). This analysis was performed using the Global Collaborative Network, which contains data from 158 HCOs with 182,329,401 patients and provides real-time access to patient demographics, diagnoses, procedures, medications, encounters, and laboratory values. Our study was conducted in accordance of the Strengthening the Reporting of Observational Studies in Epidemiology (STROBE) guidelines and followed the TriNetX publication standards. Clinical codes used for cohort definition and outcomes are provided in the supplementary table.

### Study Cohort

Two cohorts of adults aged ≥18 years with HFmrEF were constructed in TriNetX. HFmrEF was defined by a documented left ventricular ejection fraction (LVEF) between 41% and 49% and a diagnosis of HF. Eligible LVEF measurements and HF diagnosis were required to occur on or after September 1, 2021. The empagliflozin cohort included patients with newly prescribed empagliflozin, with no documented exposure to dapagliflozin. The dapagliflozin cohort included patients with newly prescribed dapagliflozin, with no documented exposure to empagliflozin.

Both cohorts were required to have no prior exposure to any SGLT2 inhibitor before the index date and had at least three documented prescriptions of the corresponding SGLT2 inhibitor. The index date was defined as the date of the first qualifying empagliflozin or dapagliflozin prescription following documentation of HFmrEF, and all inclusion criteria were required to be met on or before the index date.

### Outcomes

For each cohort, outcomes were measured one day after the index event through 365 days. Outcomes included all-cause mortality, hospitalization, urinary tract infections (UTI), and HF exacerbation. Patients with a prior history of these outcomes were excluded to assess new incidence. Rounded 365-day windows were chosen for ease of interpretation. Mortality data in TriNetX are obtained from EMRs and national death registries, with over 94% of HCOs linked to U.S. death records.

### Propensity Matching

Propensity score matching (PSM) was performed 1:1 using logistic regression and greedy nearest-neighbor matching with a tolerance of 0.1 to control for confounding variables. Matching variables included demographics (age, race, sex, and ethnicity), diagnoses, labs, and encounter characteristics. The rows of data were randomized to diminish any biases of the nearest-neighbor algorithm.

### Statistical Analysis

All statistical analyses were conducted within TriNetX using the Compare Outcomes tool. This tool provides four classes of analysis: measures of association (risk, risk difference, risk ratio, and odds ratio), survival analysis (Kaplan-Meier curves, log-rank tests, hazard ratios, and proportionality testing), number of instances analysis, and laboratory result distribution analysis. A P-value ≤0.05 was used to determine statistical significance. Comparisons were performed before and after propensity matching.

### IRB Statement

This study utilizing de-identified data from TriNetX has been determined to be exempt by the UTMB IRB. The UTMB IRB determined that this project does not involve intervention or interaction with human subjects and is de-identified per the de-identification standard defined in Section §164.514(a) of the HIPAA Privacy Rule.

## Results

### Cohort Identification and Matching

A total of 2,428 empagliflozin-treated and 2,573 dapagliflozin-treated patients met the inclusion criteria before matching. After 1:1 PSM 1,466 patients remained in each cohort (Figure 1), and no statistically significant differences were observed across baseline characteristics (Table 1).

**Figure 1.**
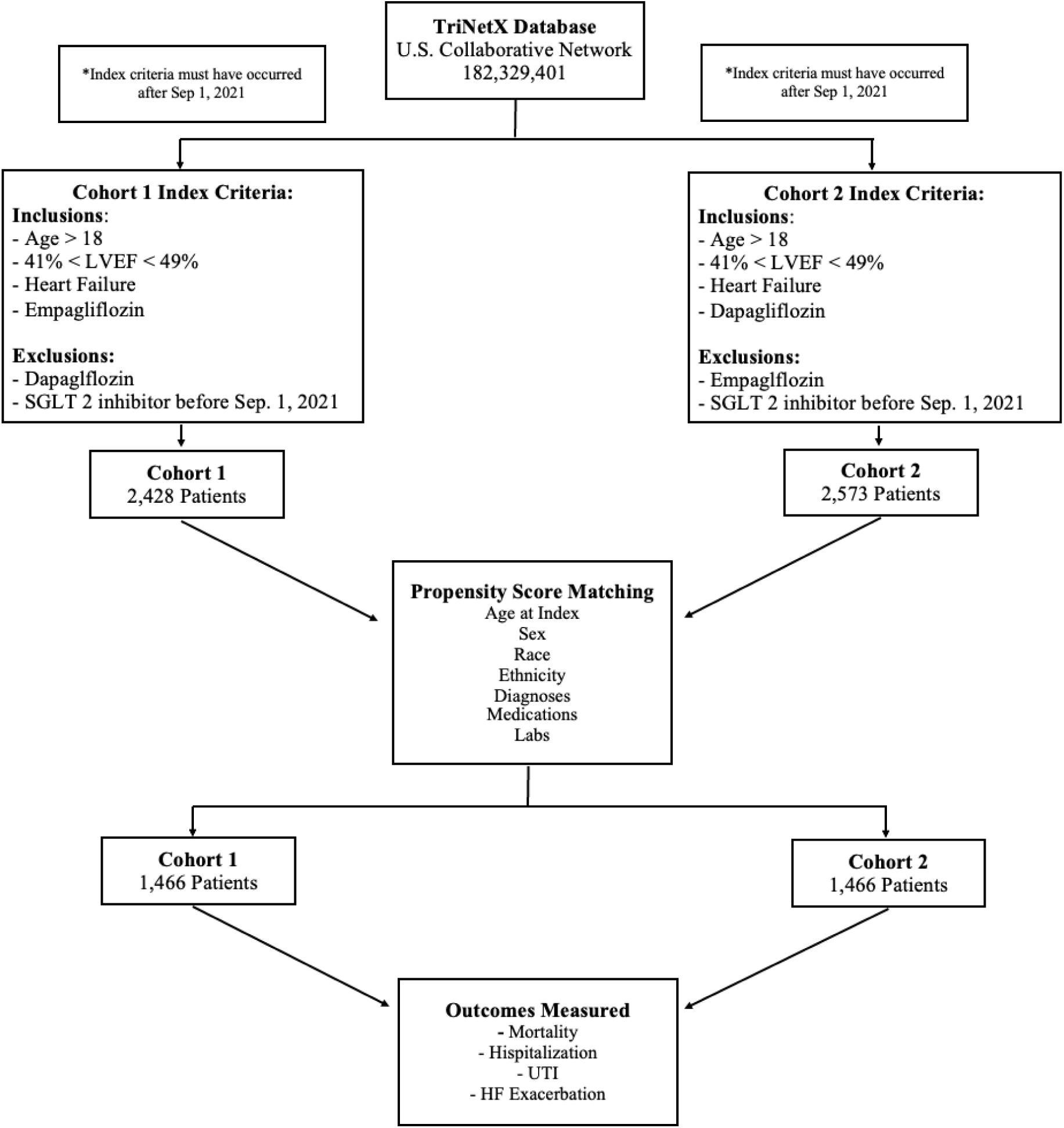
Identification of Empagliflozin and Dapagliflozin Cohorts and PSM in HFmrEF

**Table 1.** Baseline characteristics of the patient cohort before and after PSM.

|  |  |  |  |  |  |  |
| --- | --- | --- | --- | --- | --- | --- |
| **Characteristics** | **Before PSM** |  |  | **After PSM** |  |  |
|  | **Empagliflozin**<br>**(n=2428)** | **Dapagliflozin**<br>**(n=2573)** | **P-**<br>**value** | **Empagliflozin**<br>**(n=1466)** | **Dapagliflozin**<br>**(n=1466)** | **P-**<br>**value** |
| **Age at Index** |  |  |  |  |  |  |
| Mean ± SD | 67.8 ± 12.8 | 67.4 ± 13.2 | 0.221 | 67.5 ± 12.9 | 67.2 ± 13.2 | 0.503 |
| **Sex** |  |  |  |  |  |  |
| Female | 862 | 955 | <0.001 | 538 | 525 | 0.727 |
| **Ethnicity** |  |  |  |  |  |  |
| Hispanic or Latino | 155 | 175 | 0.552 | 113 | 103 | 0.48 |
| Not Hispanic or Latino | 2,012 | 2,339 | <0.001 | 1,279 | 1,305 | 0.138 |
| **Race** |  |  |  |  |  |  |
| White | 1,988 | 1,688 | <0.001 | 1,120 | 1,128 | 0.727 |
| Black or African American | 278 | 724 | <0.001 | 233 | 237 | 0.840 |
| **Diagnosis** |  |  |  |  |  |  |
| Atrial Fibrillation/Flutter | 1,149 | 1,198 | 0.589 | 678 | 667 | 0.684 |
| Systolic Heart Failure | 1,789 | 1,805 | 0.006 | 1,065 | 1,071 | 0.803 |
| Persons with Potential Health Hazards Related to Socioeconomic and Psychosocial Circumstances | 311 | 364 | 0.166 | 203 | 183 | 0.275 |
| Diastolic Heart Failure | 757 | 847 | 0.187 | 446 | 450 | 0.873 |
| Combined Diastolic and Systolic Heart Failure | 669 | 1,110 | <0.001 | 461 | 443 | 0.472 |
| Diabetes Mellitus | 1144 | 1,384 | <0.001 | 710 | 703 | 0.796 |
| **Medication** |  |  |  |  |  |  |
| Loop Diuretics | 1,629 | 1,885 | <0.001 | 990 | 982 | 0.753 |
| Beta Blockers/Related | 1,916 | 2,107 | 0.008 | 1,154 | 1,153 | 0.964 |
| Angiotensin II Inhibitors | 1,096 | 1,206 | 0.22 | 683 | 677 | 0.824 |
| ACE Inhibitors | 687 | 593 | <0.001 | 385 | 384 | 0.967 |
| Potassium Sparing/Combination Diuretics | 666 | 683 | 0.481 | 413 | 404 | 0.711 |
| Calcium Channel Blockers | 1,014 | 1,269 | <0.001 | 645 | 650 | 0.852 |
| Hydralazine | 505 | 1,156 | <0.001 | 424 | 398 | 0.285 |
| Organic Nitrates | 983 | 1,374 | <0.001 | 626 | 643 | 0.526 |
| Platelet Aggregation Inhibitors | 1,366 | 1,635 | <0.001 | 860 | 867 | 0.793 |
| Antilipemic Agents | 1,712 | 1,834 | 0.55 | 1,016 | 1,032 | 0.520 |
| Sacubitril | 407 | 518 | 0.002 | 284 | 277 | 0.742 |
| Amiodarone | 383 | 531 | <0.001 | 248 | 261 | 0.526 |
| Digoxin | 147 | 176 | 0.258 | 96 | 91 | 0.706 |
| Insulin and Analogues | 922 | 1,232 | <0.001 | 590 | 595 | 0.851 |
| Metformin | 322 | 244 | <0.001 | 161 | 157 | 0.812 |
| GLP-1 Analogues | 144 | 75 | <0.001 | 56 | 59 | 0.775 |
| DPP-4 Analogues | 47 | 66 | 0.134 | 35 | 33 | 1 |
| Sulfonylureas | 114 | 95 | 0.076 | 62 | 59 | 0.781 |
| **Labs** |  |  |  |  |  |  |
| GFR | 70.4 ± 29.2 | 70.1 ± 31.3 | 0.733 | 69.8 ± 29.6 | 70.2 ± 30.2 | 0.756 |
| Hemoglobin A1c | 6.8 ± 1.7 | 6.9 ± 1.9 | 0.027 | 6.8 ± 1.8 | 6.8 ± 1.8 | 0.783 |
| BNP | 2890 ± 5626 | 946 ± 1714 | <0.001 | 2996 ± 6235 | 928 ± 1780 | <0.001 |
| BNP Prohormone | 3909 ± 6054 | 5687 ± 8379 | <0.001 | 3672 ± 5157 | 5329 ± 8110 | <0.001 |
| LVEF | 43.4 ± 8.9 | 41.5 ± 9.8 | <0.001 | 43.2 ± 9.4 | 42.7 ± 8.8 | 0.141 |
| Abbreviations: PSM, propensity score matching; ACE, angiotensin converting enzyme; GLP, glucagon-like-peptide; DPP, dipeptidyl peptidase; GFR, glomerular filtration rate; BNP, B-type natriuretic peptide; LVEF, left ventricular ejection fraction |  |  |  |  |  |  |

### Baseline Characteristics

Prior to matching, several clinical, demographic, and treatment-related differences were observed (Table 1). The dapagliflozin cohort had higher representation of black patients, systolic HF, and combined systolic/diastolic HF. Differences were also present in multiple cardiovascular therapies, including loop diuretics, beta blockers, angiotensin receptor blockers, calcium channel blockers, hydralazine, organic nitrates, antiplatelet agents, and lipid-lowering agents. Significant laboratory differences were present for BNP, BNP prohormones, and LVEF.

After PSM, all demographic variables, HF subtypes, comorbid conditions, medication classes, and laboratory parameters were well balanced without statistically significant differences (Table 1). This included similar distributions of atrial fibrillation/flutter, systolic and diastolic HF, diabetes mellitus, psychosocial health-hazard codes, cardiometabolic therapies, natriuretic peptides, and LVEF. Matched cohorts also demonstrated comparable utilization of all cardiovascular and diabetes-related therapies, including diuretic classes, renin-angiotensin-aldosterone system modulators, hydralazine, antiplatelet agents, and insulin-based therapies.

### Clinical Outcomes

In the propensity-matched cohort, all-cause mortality did not differ significantly between treatment groups (Table 2). Mortality occurred in 7.9% of empagliflozin-treated patients and 9.8% of dapagliflozin-treated patients, corresponding to an absolute risk difference of –1.9% (95% CI –4.0% to 0.1%, P=0.069) in Table 2. Time-to-event analysis demonstrated a significantly lower hazard of mortality in the empagliflozin cohort (HR 0.75, 95% CI 0.59-0.96), with Kaplan-Meier curves separating early and remaining distinct throughout follow-up (log-rank χ² =5.28, P=0.022) (Table 2, Figure 2A).

**Figure 2:**
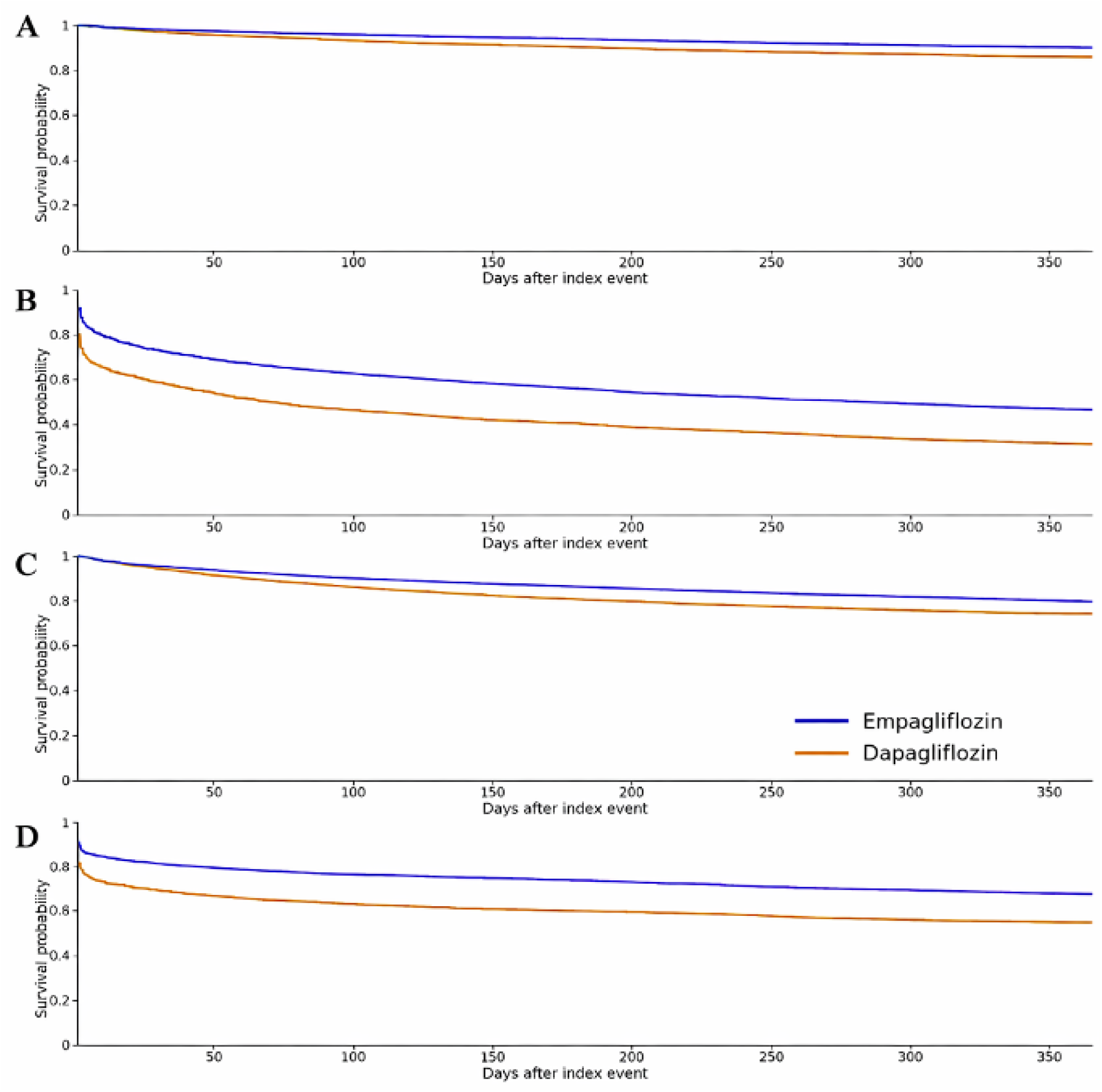
Kaplan-Meier curves for mortality (A), hospitalization (B), urinary tract infection (C), and heart failure exacerbation (D) following propensity score matching. Blue lines denote empagliflozin and orange lines denote dapagliflozin. Group differences were assessed by log-rank testing reported in Table 2.

**Table 2.**
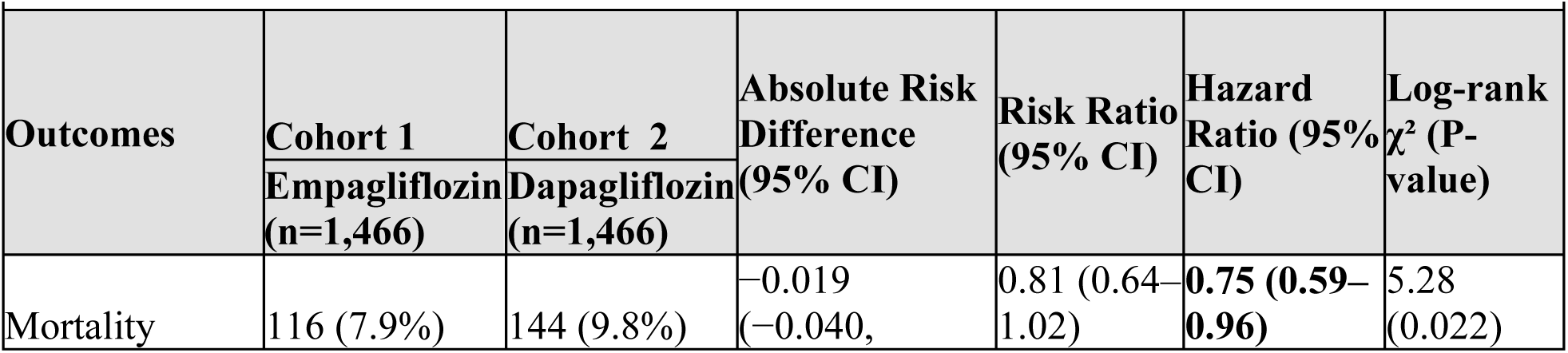
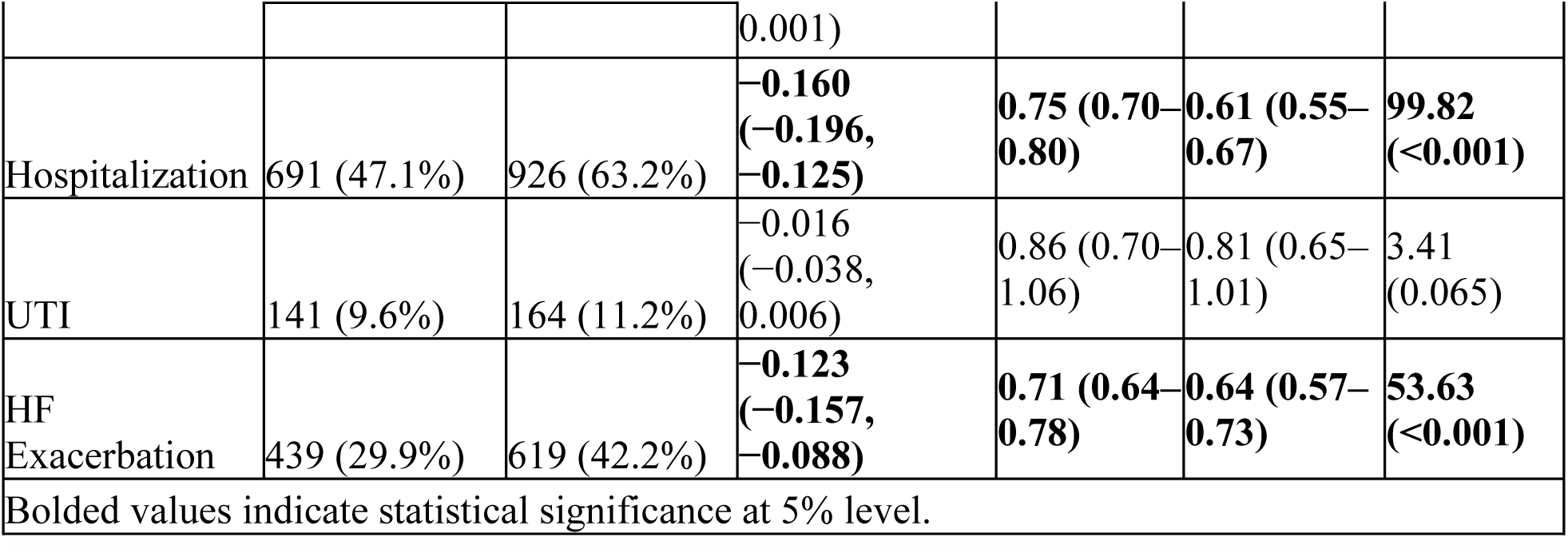
Risk Difference between outcomes of interest between Empagliflozin and Dapagliflozin Cohorts After PSM.

Hospitalization occurred significantly less frequently among empagliflozin-treated patients compared with dapagliflozin-treated patients (Table 2). Event rates were 47.1% vs 63.2%, respectively, yielding an absolute risk reduction of 16.0% (95% CI –19.6% to –12.5%, P<0.001) and a risk ratio of 0.75 (95% CI 0.70-0.80). Survival analysis further demonstrated a 39% lower hazard of hospitalization with empagliflozin (HR 0.61, 95% CI 0.55-0.67), supported by a highly significant log-rank test (χ² = 99.82, P<0.001) (Table 2, Figure 2B). Additionally, empagliflozin-treated patients experienced fewer recurrent hospitalizations, with a lower mean number of events during follow-up (P=0.018).

Rates of UTI were similar between groups, occurring in 9.6% of empagliflozin-treated and 11.2% of dapagliflozin-treated patients (Table 2). The absolute risk difference was –1.6% (95% CI –3.8% to 0.6%, P=0.164), with no significant difference in relative risk (RR 0.86, 95% CI 0.70-1.06). Time-to-event and recurrent-event analyses likewise showed no significant differences, with overlapping Kaplan-Meier curves and a non-significant hazard ratio (HR 0.81, 95% CI 0.65-1.01, log-rank **χ²**=3.41, P=0.065) (Table 2, Figure 2C).

Empagliflozin treatment was associated with a significantly lower risk of HF exacerbation compared with dapagliflozin (Table 2). Exacerbations occurred in 29.9% of empagliflozin-treated patients, corresponding to an absolute risk reduction of 12.3% (95% CI –15.7% to –8.8%, P<0.001) and a risk ratio of 0.71 (95% CI 0.64-0.78). Kaplan-Meier analysis confirmed a 36% lower hazard of HF exacerbation with empagliflozin (HR 0.64, 95% CI 0.57-0.73), with significant separation of survival curves (log-rank χ² =53.63, P<0.001) (Table 2, Figure 2D). Rates of recurrent exacerbations did not differ significantly between groups.

## Discussion

In this large, real-world cohort of patients with HFmrEF, treatment with empagliflozin was associated with a lower all-cause mortality, fewer hospitalizations, and reduced HF exacerbations compared with dapagliflozin over one year of follow-up. Rates of UTI were similar between groups, supporting comparable class-related safety. These findings suggest potential heterogeneity in clinical outcomes among individual agents within the SGLT2 inhibitor class. Further investigation is warranted to elucidate the mechanisms underlying these observed differences.

When interpreted in the context of existing real-world evidence, the findings of this study suggest that conclusions drawn from broader HF populations may not fully capture treatment effects within specific phenotypes. Prior comparative analyses of empagliflozin and dapagliflozin have generally reported comparable outcomes when evaluated across aggregated EF categories, supporting class-level effectiveness of SGLT2 inhibitors^3,7^. However, such approaches may dilute clinically relevant differences that emerge within more narrowly defined populations. By isolating patients with HFmrEF, the present analysis reveals outcome differences between empagliflozin and dapagliflozin that are not readily apparent in broader comparisons. These findings underscore the importance of phenotype-specific evaluation and suggest that therapeutic equivalence observed at the population level may not be uniformly applied across all HF subgroups.

SGLT2 inhibitors have emerged as foundational therapy for HF, demonstrating consistent reductions in HF-related events across the spectrum of LVEF. The cardiovascular benefits of this class were first established in EMPA-REG OUTCOME, which showed significant reductions in cardiovascular death, HF hospitalization, and all-cause mortality among patients with T2DM treated with empagliflozin ^8^. Beyond glycemic control, SGLT2 inhibitors are believed to reduce cardiovascular events through multiple mechanisms, including inhibition of cardiac sodium transporters, attenuation of inflammation and oxidative stress, and improvements in myocardial energetics and ventricular loading conditions, collectively contributing to enhance cardiac function and reduced HF symptoms ^9^. Additional mechanistic studies suggest that SGLT2 inhibitors may also improve endothelial function and reduce myocardial oxidative stress, further supporting their role in modifying HF pathophysiology across diverse clinical phenotypes ^10,11^.

Although dapagliflozin and empagliflozin share a common mechanism of renal SGLT2 inhibition, pharmacokinetic and pharmacodynamic differences between agents have been reported. Empagliflozin has been shown to exhibit greater potency, a longer elimination half-life, and enhanced bioavailability compared with dapagliflozin, which may contribute to differential tissue-level effects and provide a biologically plausible rationale for differences in clinical outcomes ^6,12^. At the same time, observational studies and pooled analyses have shown similar HF and renal benefits with empagliflozin and dapagliflozin, supporting strong class-wide effects while allowing for potential agent-level differences in selected populations ^13^.

Randomized outcome trials evaluating empagliflozin in HF have primarily focused on populations with reduced or preserved EF. The EMPEROR-Reduced trial demonstrated a significant reduction in the composite of cardiovascular death or HF hospitalization in patients with an EF ≤40%, although it was not powered to detect cardiovascular mortality alone ^14^.

Similarly, the EMPEROR-Preserved trial extended these findings to patients with EF >40%, with benefits driven primarily by reductions in HF hospitalization rather than mortality ^2^.

Importantly, neither trial was designed to specifically evaluate outcomes in patients with HFmrEF, despite emerging evidence that SGLT2 inhibitors may prevent or reverse adverse cardiac structural remodeling during this transitional stage of HF ^6,15^. Meta-analytic data incorporating DAPA-HF and EMPEROR-Reduced further support reductions in cardiovascular death and HF hospitalization with SGLT2 inhibition in reduced EF populations, providing a broader framework for extending benefit across the EF continuum ^16^.

Similarly, the DELIVER trial evaluated dapagliflozin in patients with HF and EF>40%, including those with recovered EF following prior systolic dysfunction. Although dapagliflozin significantly reduced worsening HF events, cardiovascular mortality was not significantly reduced, and observed benefits were largely driven by reductions in hospitalization ^5^. Differences in study design, including inclusion of patients with improved EF, shorter follow-up duration, and heterogeneity in background medical therapy, may have limited the ability to detect mortality effects. Real-world comparative analyses of empagliflozin and dapagliflozin have likewise reported broadly similar HF outcomes, underscoring ongoing uncertainty regarding agent-specific differences and the need for focused studies in HFmrEF populations ^17^.

Collectively, these trials demonstrate consistent reductions in HF worsening across EF categories but provide limited evidence for agent-specific or mortality-driven benefits, particularly in patients with HFmrEF. In contrast, the present study uniquely provides a head-to-head comparison of empagliflozin and dapagliflozin in a well-defined HFmrEF population, excluding patients with prior SGLT2 inhibitor exposure. By leveraging a large, heterogeneous real-world cohort and applying rigorous PSM, this analysis complements randomized trial data and offers clinically relevant insights into comparative effectiveness. The findings suggest that empagliflozin may be associated with more favorable mortality and morbidity outcomes than dapagliflozin in HFmrEF.

### Limitations

Several limitations of this study should be acknowledged. Although empagliflozin and dapagliflozin are prescribed at standardized doses for HF, real-world prescribing patterns may include higher doses, particularly when used for glycemic management. Prior comparative effectiveness research has suggested potential dose-dependent differences in outcomes, however, reliable dose-stratified analyses are not feasible within the TriNetX platform^18^. As such, differences in dosing could not be evaluated in the present study and may contribute to residual heterogeneity. Future prospective studies or datasets with granular medication dosing information are needed to clarify the clinical implications of SGLT2 inhibitor dose variation in patients with HF. The retrospective observational design precludes causal inference. Although associations between empagliflozin and improved clinical outcomes were observed, treatment assignment was not randomized, and residual confounding cannot be fully excluded. Extensive propensity score matching was performed to balance demographics, comorbid conditions, medication use, and laboratory parameters between treatment groups, however, unmeasured or incompletely captured variables may have influenced outcomes. Factors such as HF severity beyond recorded EF, duration of disease, medication adherence, socioeconomic status, and provider-level prescribing preferences were not available for matching and may have contributed to residual bias. Even with well-balanced matched cohorts, unobserved confounding remains an inherent limitation of real-world analyses. This analysis was conducted using de-identified data from the TriNetX Global Collaborative Network, which limited the ability to evaluate clustering effects at the level of individual hospitals or health systems. Institutional practice patterns, regional differences in care delivery, and variations in access to specialized HF management could not be assessed due to data privacy restrictions, precluding evaluation of site-level heterogeneity and its potential impact on outcomes. Follow-up was limited to one year after treatment initiation, and longer-term comparative outcomes, including durability of benefit and late adverse events, were not assessed and warranted further investigation in studies with extended follow-up.

## Conclusion

In this large, real-world propensity-matched analysis of patients with HFmrEF, empagliflozin was associated with lower hazards of all-cause mortality, hospitalization, and HF exacerbation compared with dapagliflozin, with no significant differences in safety outcomes. These findings suggest that clinical effectiveness may vary within the SGLT2 inhibitor class and further support HFmrEF as a distinct and clinically relevant HF phenotype. Prospective, adequately powered comparative studies are needed to validate these observations and to determine whether pharmacologic differences among individual agents result in meaningful differences in long-term cardiovascular outcomes.

## Sources of Funding

This research was supported by the UTMB Institute for Translational Sciences, supported in part by a Clinical and Translational Science Award (UL1 TR001439) from the National Center for Advancing Translational Sciences at the National Institutes of Health (NIH). The content is solely the responsibility of the authors and does not necessarily represent the official views of the NIH.

## Disclosures

The authors have no competing interests to declare that are relevant to the content of this article.

## Data Availability

All data are publicly available in the TriNetX Database through institutional agreement.

https://live.trinetx.com/

---

**Supplemental Table S1.**
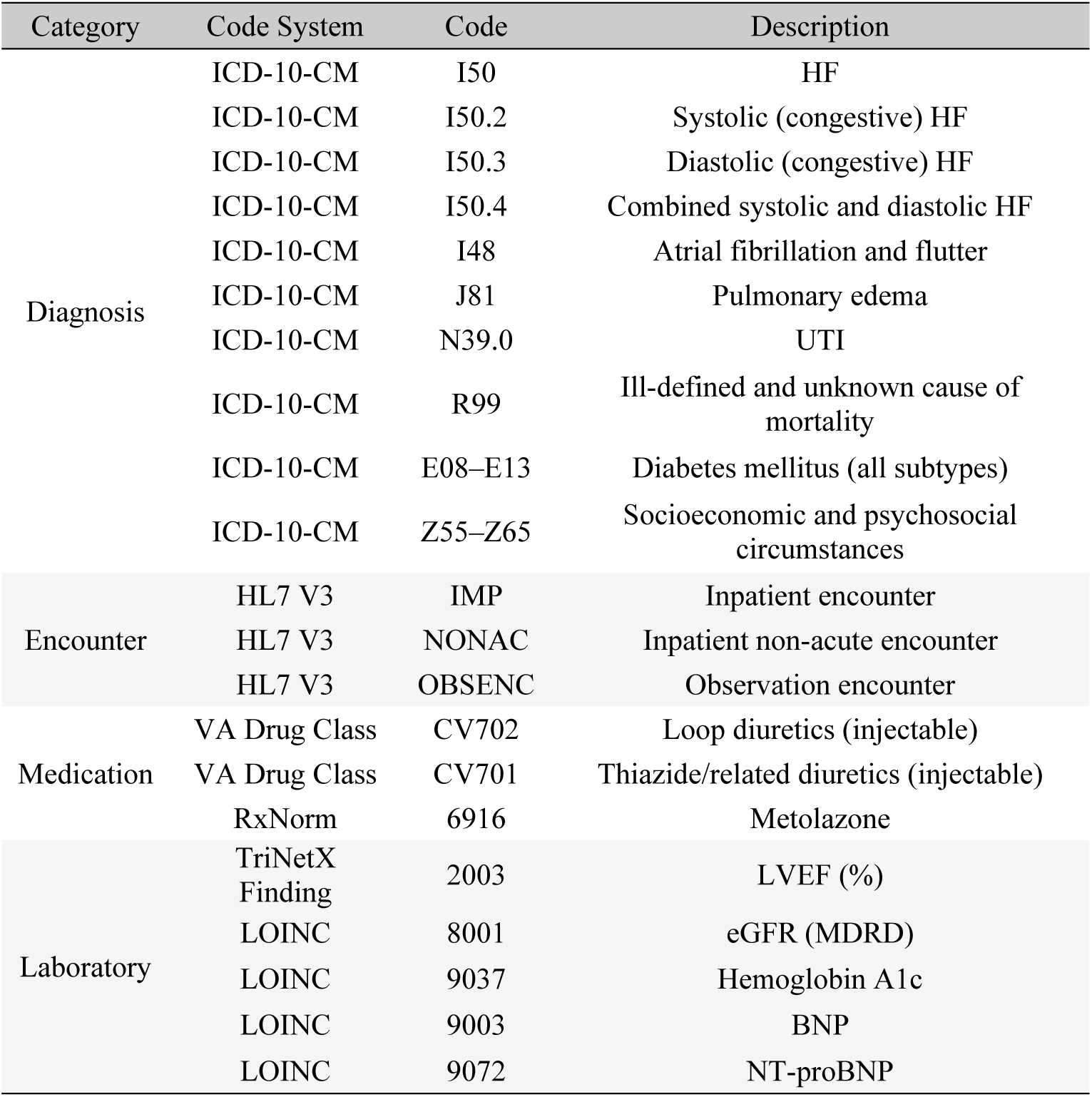
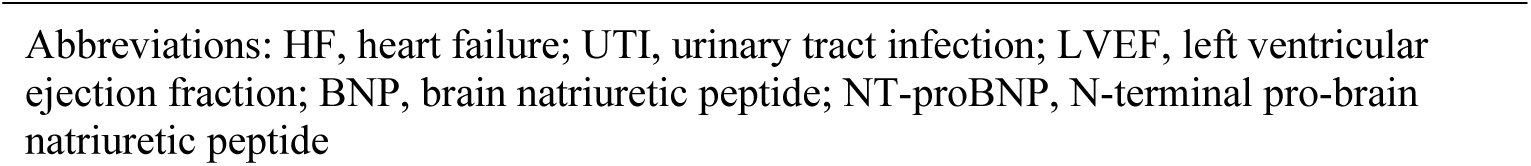
Clinical Codes Used for Cohort Definition and Outcomes

## Notes

### Competing Interest Statement

The authors have declared no competing interest.

### Author Declarations

This study utilizing de-identified data from TriNetX has been determined to be exempt by the UTMB IRB. The UTMB IRB determined that this project does not involve intervention or interaction with human subjects and is de-identified per the de-identification standard defined in Section 164.514(a) of the HIPAA Privacy Rule.

